# The long-term effects of free care on birth outcomes: Evidence from a national policy reform in Zambia

**DOI:** 10.1101/2021.05.18.21257410

**Authors:** Mylene Lagarde, Aurelia Lepine, Collins Chansa

## Abstract

As women in many countries still fail to give birth in facilities due to financial barriers, many see the abolition of user fees as a key step on the path towards universal coverage. We exploited the staggered removal of user charges in Zambia from 2006 to estimate the effect of user fee removal over up to five years after the policy change. We used data from the birth histories of two nationally representative Demographic and Health Surveys to implement a difference-in-differences analysis and identify the causal impact of removing user charges on institutional and assisted deliveries, caesarean sections and neonatal deaths. We also used the rich survey data to explore heterogeneous effects of the policy. Removing fees had little effect in the short term but large positive effects appeared about two years after the policy change. Institutional deliveries in treated areas increased by 25 to 35%, driven entirely by a reduction in home births. However, there was no evidence that the reform changed the behaviours of women with lower education, the proportion of caesarean sections or reduced neonatal mortality. Institutional deliveries increased where care quality was high, but not where it was low. While abolishing user charges may reduce financial hardship from healthcare payments, it does not necessarily improve equitable access to care or health outcomes. Shifting away from user fees is a necessary but insufficient step towards universal health coverage, and concurrent reforms are needed to target vulnerable populations and improve quality of care.

## 1. Introduction

In 2017, an estimated 300,000 women died during or following pregnancy and child birth, while 2.5 million children died in the first month of life in 2018, mostly due to issues arising at birth or immediately after (WHO 2020). The vast majority of these deaths could be avoided with better access to cost-effective interventions and skilled care (Horton and Levin 2016), but many women still fail to deliver in facilities. Although low use of modern healthcare services is caused by a range of factors, such as limited accessibility, poor quality or cultural barriers (Gage 2007), lack of money remains one of the most significant barriers to accessing care, especially when health facilities charge for providing care at the point of delivery, which still occur in many low- and middle-income countries (Saksena, Xu et al. 2010). As a result, many see the abolition of user fees as a key step on the path towards universal coverage – defined as ensuring timely access to quality healthcare without financial hardship (WHO 2010).

Despite the high policy relevance of this reform, and the heated debate on user fees more generally, there has been a relative dearth of rigorous empirical evidence on the impact of user charges. Several randomised trials have looked at the effect of price on the uptake of preventive health care products (e.g. bednets, deworming drugs, chlorine) and concluded that even small nominal fees could have large negative deterrent effects on utilisation (Dupas and Miguel 2017). Randomised trials in Mali and Ghana have also found benefits of free care on health service use, and even some improvement in health outcomes (Ansah, Narh-Bana et al. 2009, Powell-Jackson, Hanson et al. 2014, Sautmann, Brown et al. 2020). Yet the narrow scope of these trials, either focused on specific products or a few facilities, limits the extent to which one can generalise lessons to system-wide health financing reforms. In practice, the positive effects of free care can be thwarted by issues that often plague complex health system reforms. First, if the abolition of user fees is imperfect (i.e. fees are still charged), individuals have no reason to change their behaviours. Second, if removing fees fuels a deterioration of quality of care, through drug stock-outs or increased absenteeism of demotivated staff, individuals may be discouraged to use health services. If they still decide to use services, poor quality of care may limit the extent to which increased use translates into improved health outcomes.

Several studies have sought to shed a light on the effects of abolishing user fees at scale, but none has looked at the long-term effects on maternal care-seeking or health outcomes. Observational studies have often failed to identify the causal effects of abolishing user fees, or have been limited by the use of data from facility registers – that can be unreliable and restrict what researchers can study (Lagarde and Palmer 2008, Dzakpasu, Powell-Jackson et al. 2014). Studies that have used more robust data and statistical approaches have pointed to mixed effects of removing fees on care-seeking for acute illnesses (Hangoma, Robberstad et al. 2018, Lepine, Lagarde et al. 2018) Recent studies have tried to shed a light on the benefits of removing fees on maternal care seeking, by comparing areas (or countries) that removed fees to others that did not (McKinnon, Harper et al. 2014, Chama-Chiliba and Koch 2016, Leone, Cetorelli et al. 2016). The evidence of effects is mixed, with a previous analysis of the reform in Zambia showing no change in facility-based deliveries (Chama-Chiliba and Koch 2016). Existing studies have not explored how the effect of removing fees evolves over time, even though beneficial effects may take time to appear in the short-term due to teething problems or slow behavioural change (Carasso, Lagarde et al. 2012).

When Zambia initially introduced user fees in 1991, its objectives were similar to many other sub-Saharan African countries: raising additional income to improve the quality of services, and improving staff motivation and accountability through community participation and salary top-ups (Government of the Republic of Zambia 1991). In theory, exemptions were in place for certain groups of the population (e.g. children under 5 year old, indigents) or services (antenatal services) (Chama-Chiliba and Koch 2016). In practice, fee exemptions were poorly enforced, leading to inequitable access to health services (Cheelo, Chama et al. 2010). In light of such negative aspects, user fees were removed from April 2006 in the 54 rural districts of the country. Fees remained in place in the other 18 districts, until the free care policy was extended to the peri-urban parts of these districts in June 2007, and then to the entire country in 2012.

In this study, we aimed to examine the causal effect of removing user charges on birth outcomes, using the staggered implementation of the policy change in 2006 and 2007. We used data from two nationally representative surveys and implemented a difference-in-differences strategy to evaluate the impact of removing fees up to five years after the policy reform on the place and type of delivery, as well as neonatal mortality.

## 2. Methods

### Data

We used information contained in the 2007 and the 2013-2014 Zambia Demographic and Health Surveys (DHS) for all of the births that women had in the five years before the interview. These birth histories allowed us to construct a dataset containing detailed information on births over a ten-year period spanning over the two policy changes (see the timeline of policy reforms and data used in Figure B1 in Online appendix).

Given the staggered rollout of the policy, we defined three groups in our datasets: (1) individuals living in rural districts where care was free from April 1st 2006; (2) peri-urban areas in urban districts where fees are removed on June 1st 2007 and (3) urban areas of urban districts where fees remained in place until January 2012. For the purpose of our analysis we excluded any birth that occurred after January 2012, so that urban areas remain a control group where user charges apply throughout the analysis period.

For each birth, we considered the effect of the policy on four outcomes.

Firstly, we considered the place of delivery, and constructed a binary indicator equal to 1 if the woman gave birth in a public or mission healthcare facility where the policy change occurred, and 0 otherwise. Secondly, we considered if the woman was assisted by a skilled birth attendant (doctor, nurse or midwife) during a delivery. This is relevant because maternal and neonatal health outcomes are likely to be better in the presence of a qualified staff. In addition, if the policy change fuelled staff shortages, the proportion of assisted deliveries could have fallen.

Thirdly, we considered whether the delivery was done by caesarean section, as an increase in the volume of deliveries without an adequate response of the supply side could lead to a reduction in the proportion of C-sections undertaken. Finally, we considered neonatal mortality, specifically whether the child dies on the day of the delivery or within the first 28 days. Both are strongly linked to the conditions in which women deliver, and can be seen as potential indicators of the effectiveness of care received.

Table 1 shows the descriptive statistics for the analytical sample of mothers and births, spanning the period 2002-2011. A few salient facts should be noted between the two sets of ‘treated’ areas where fees were removed (rural districts and peri-urban areas) and the control (urban) areas. In treated areas, women were from less wealthy households, had more children and were more likely to have a lower education level. There were also fewer institutional deliveries in these treated areas compared to urban areas, although the overwhelming majority of these deliveries were assisted by a qualified staff. Only a small proportions of births were done by caesarean sections (from 3% in rural and peri-urban areas to 7% in urban areas), and less than 3% of babies born died within the first 28 days.

**Table 1.** Sample description.

|  | **Rural districts** | **Peri-urban areas** | **Urban areas** |
| --- | --- | --- | --- |
| **Mothers** |  |  |  |
| Age (years) | 29.52 (7.17) | 29.03 (7.12) | 28.53 (6.63) |
| Wealth index | -0.21 (0.79) | -0.25 (0.74) | 0.83 (1.30) |
| Number of children | 4.30 (2.52) | 4.26 (2.57) | 3.37 (2.20) |
| Has no education or incomplete primary | 4269 (59%) | 711 (60%) | 810 (29%) |
| **Births** |  |  |  |
| Institutional deliveries | 4974 (53%) | 742 (45%) | 2839 (81%) |
| Home births | 4430 (47%) | 887 (54%) | 639 (18%) |
| Assisted deliveries | 4649 (49%) | 677 (41%) | 2795 (80%) |
| Caesarean sections | 252 (3%) | 50 (3%) | 246 (7%) |
| Neonatal deaths | 260 (3%) | 38 (2%) | 103 (3%) |

Data are n (%) or mean (SD).

### Statistical analysis

We use a Difference-in-difference (DiD) approach to identify the intention-to-treat (ITT) effect of the policy. For a given birth event *y*_*idt*_ occurring at time *t* for individual *i* living in district *d*, we estimated a specification of the form:

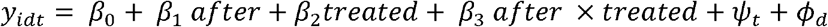

where the variable *after* is coded 1 if the child was delivered after user fees were removed and 0 otherwise and *treated* is a dummy variable coded 1 if the woman currently lives in a treated area, and 0 if she lives in a control area. We also include district (*ϕ*_*d*_) and year (*ψ*_*t*_) fixed effects. The ITT effect of user fees removal on outcome is given by 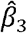 on the interaction term. Although all outcomes are binary, we estimated linear regressions for ease of interpretation so that 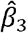 can be interpreted as the (percentage point) increase in the outcome in the treated group, compared to its pre-reform level. We also present results from logistic regressions in the online Appendix. We undertook two separate analyses. First, to identify the effect of the policy of the first phase of the policy roll out occurring in April 2006, we restricted the sample to rural districts (treated areas) and urban areas of urban districts (control areas). This analysis estimated the effect of the policy in rural districts. Second, we identified the effect of the policy in peri-urban areas, and restricted the sample to peri-urban (treated) and urban (control) areas of urban districts, with the policy change occurring from June 2007.

We used the location of the DHS sampling cluster in which a woman lived at the time of the interview to infer which policy has applied to her during her entire birth history. As DHS data include the name of the district in which the household lives, determining which women lived in one of the 54 districts in which the 2006 policy change occurred was straightforward. To determine whether a woman lived in peri-urban areas of urban districts, we used the GIS coordinates of her sampling cluster and calculated the distance to the administrative centre of the district, the criteria used by health authorities to identify peri-urban areas – see Appendix C in the Online appendix for further details. Note that the assignment to treatment status makes two assumptions. First, we assumed that a woman has always resided in the same area in the last five years. Second, we assumed that the random displacement of the DHS sampling clusters does not interfere with assignment with the treatment status (Perez-Heydrich, Warren et al. 2013). We discuss these assumptions later.

A key identifying assumption for a valid DiD estimation is that outcomes in the treatment and control group were following a similar path before the policy change. We provide graphical evidence to check this assumption in Figures B2-B4 in the online appendix. The data support the assumption for most outcomes, except neonatal mortality, where trends are only parallel from 2003. Hence, we excluded data from 2002 for this outcome.

Beyond the analysis of the main effects of the policy change, we performed three sub-group analyses. To avoid performing an under-powered analysis, we do not perform this analysis on the two outcomes linked to more rare events (caesarean sections and neonatal deaths). First, we considered whether the policy benefitted differently women coming from the poorest and richest households (see supplementary material for the definition of the wealth quintiles). Second, we looked at the effects for women with low education (no or incomplete primary education) and others. Another important policy question, less frequently studied in the literature on fee removal is whether the quality of care provided in a facility contributed to women’s decisions to give birth in a facility, and to changes in health outcomes. We looked at the effects of the policy in areas with low or high quality care at the time of the delivery, based on a proxy indicator for care quality defined based on the average quality of antenatal care received by women in the area (see section D of the online Appendix for more details).

## 3. Results

Table 2 presents the main results of the difference-in-difference analysis, which can be interpreted as the average effect of the policy change (results from logistic regressions are in Table A1 in the online Appendix). Looking at the first panel of the table, the results indicate an increase in the probability to deliver in a facility by 15 percentage points in rural districts over the 2006-2011 period (95%CI: 0.11 to 0.19, p<0.0001), and 10 percentage points in peri-urban areas between 2007-2011 (0.04-0.17, p=0.001). Compared to a pre-reform proportion of 39.9% in rural districts and 37.6% in peri-urban areas, this corresponds to an increase in institutional deliveries of 35% and 25% respectively. In a complementary analysis (see Table A2 in online appendix), we show that this increase is entirely driven by a substantial reduction in home births, and not to a substitution away from deliveries in private facilities, which are relatively rare. Results from the second panel provide reassuring evidence that the increased utilisation did not reduce the proportion of assisted deliveries, as increases in this outcome are of the same magnitude as the increase in institutional delivery. There was an increase by 12 percentage points (0.07-0.16, p<0.0001) in rural districts and 8 percentage points in peri-urban areas (0.02-0.14, p=0.012). A key question is whether institutional deliveries improved health outcomes for new-borns. We find no evidence that the increase in institutional deliveries translated into a reduction in neonatal deaths (p=0.570 in rural districts and p=0.821 in peri-urban areas). Similarly, there is no evidence that the policy had an effect on the proportion of deliveries by caesarean sections (p=0.457 in rural districts and p=0.570 in peri-urban areas).

**Table 2.** Effects of user fee removal.

|  | **Policy change in rural districts** |  | **Policy change in peri-urban areas** |  |
| --- | --- | --- | --- | --- |
|  | Coefficient (95%CI) | p-value | Coefficient (95%CI) | p-value |
| **Delivery in an institutional facility** |  |  |  |  |
| Policy effect | 0.15 (0.11 to 0.19) | <0.0001 | 0.10 (0.04 to 0.17) | 0.001 |
| Mean pre-reform in 'treated' group | 0.40 |  | 0.38 |  |
| N | 12927 |  | 5126 |  |
| R<sup>2</sup> | 0.19 |  | 0.24 |  |
| **Assisted delivery at birth** |  |  |  |  |
| Policy effect | 0.12 (0.07 to 0.16) | <0.0001 | 0.08 (0.02 to 0.14) | 0.012 |
| Mean pre-reform in 'treated' group | 0.39 |  | 0.34 |  |
| N | 12910 |  | 5119 |  |
| R<sup>2</sup> | 0.19 |  | 0.25 |  |
| **Delivery by Caesarean section** |  |  |  |  |
| Policy effect | 0.01 (-0.02 to 0.04) | 0.457 | 0.01 (-0.02 to 0.04) | 0.570 |
| Mean pre-reform in 'treated' group | 0.01 |  | 0.02 |  |
| N | 12948 |  | 5132 |  |
| R<sup>2</sup> | 0.03 |  | 0.02 |  |
| **Neonatal deaths** |  |  |  |  |
| Policy effect | 0.00 (-0.01 to 0.02) | 0.570 | -0.00 (-0.02 to 0.02) | 0.821 |
| Mean pre-reform in 'treated' group | 0.03 |  | 0.03 |  |
| N | 12980 |  | 5144 |  |
| R<sup>2</sup> | 0.01 |  | 0.01 |  |

Notes: Each coefficient comes from an OLS regression that includes year and district fixed effects. Standard errors are clustered at mother level, sampling weights included.

Figure 1 presents the effects of removing fees over the years following the policy change, for the four main outcomes. Figure 1A shows the gradual effects of the policy in rural districts while Figure 1B shows the results for peri-urban areas. The results in both settings are consistent with the idea that it takes some time for women to change their behaviour and start giving birth in facilities. In rural districts, the positive effects of the policy on institutional and assisted deliveries only start to kick off three years after its implementation (the confidence intervals in 2008 are quite large probably due to the limited number of observations, but the point estimate suggests a positive effect). In peri-urban areas, the positive effect appears more quickly, one and a half years after the initial roll-out in June 2017, possibly due to the early lessons gained from the implementation in rural areas. The findings also confirm the absence of effect on caesarean sections and neonatal deaths, with the exception of a small increase in C-sections in 2008 in rural areas, which appears to be a fluke as it does not persist after.

**Figure 1.**
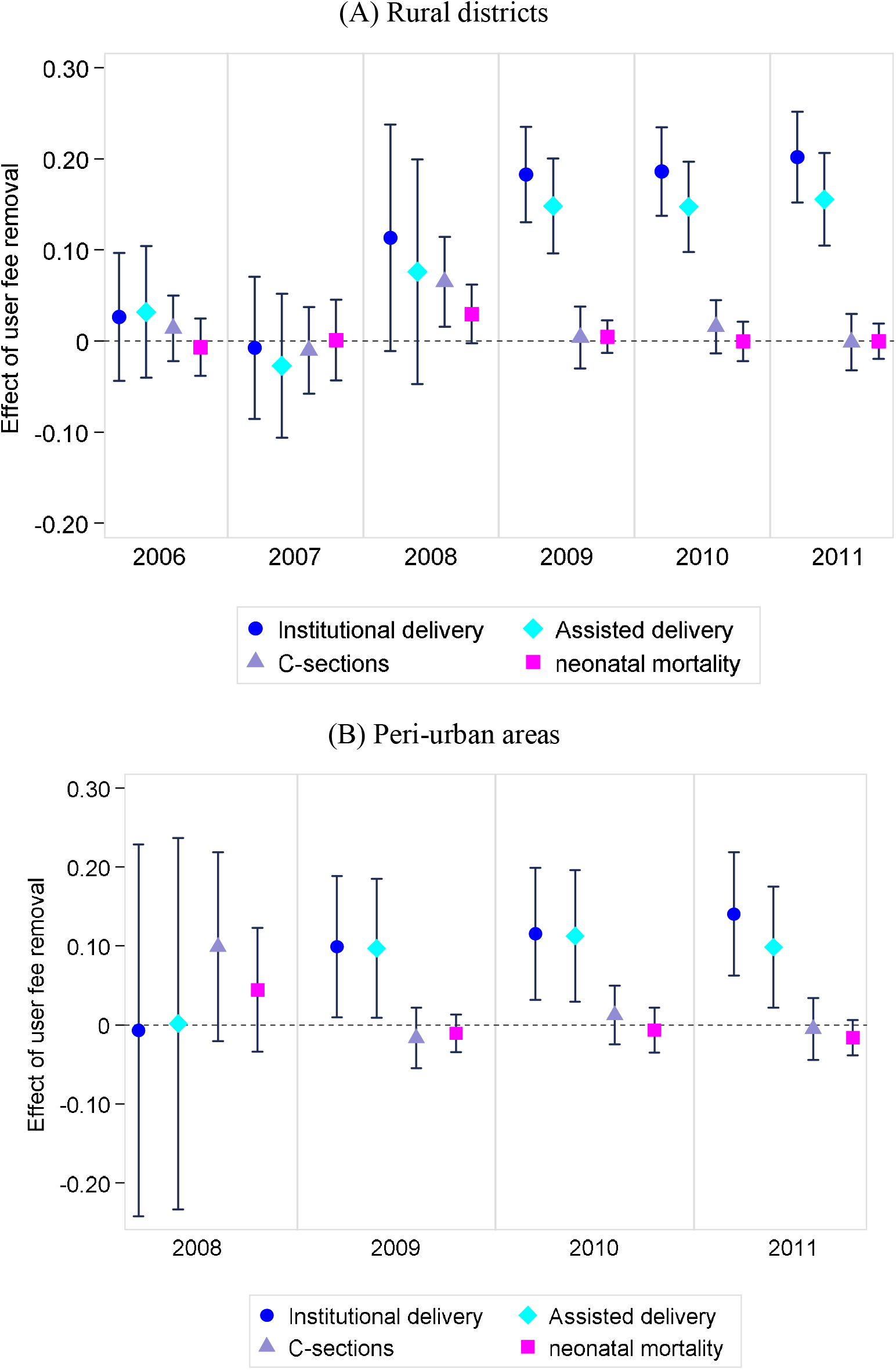
Effect of user fees removal on deliveries in institutional delivery over time. Note: The effect of the policy is represented for each year, with its confidence intervals. Each effect is estimated by using district and year fixed effects. Standard errors are clustered at the mother level, sampling weights included. Note that in figure 2B, the effect for 2007 is not presented given that only 10 children were born after June in a peri-urban areas.

Results of the sub-group analyses are presented in Figure 2A (rural districts) and 2B (peri-urban areas), while the corresponding results are in the online Appendix (Table A3). Three results emerge. First, women from the poorest quintiles benefited directly from user fee removal, with an 18 percentage points increase in institutional deliveries in rural districts and 22 percentage points in peri-urban areas. By contrast, there was no change in the choices of women from the richest quintile in either group, as most women in these groups were already delivering in facilities before the policy change (76% in rural districts and 80% in peri-urban areas). Second, despite these encouraging effects about the benefits for women from the poorest quintiles, women with lower education did not deliver more in facilities after the policy chance. In both rural and peri-urban areas, user fee removal led to a large increase in the likelihood of delivery in facilities (assisted and not) for women completing at least primary education, but not much for those with no or incomplete primary education. Third, the policy change had no effect on the proportion of institutional deliveries in areas with the lowest quality of care, while the impact of positive in areas with the highest quality care.

**Figure 2.**
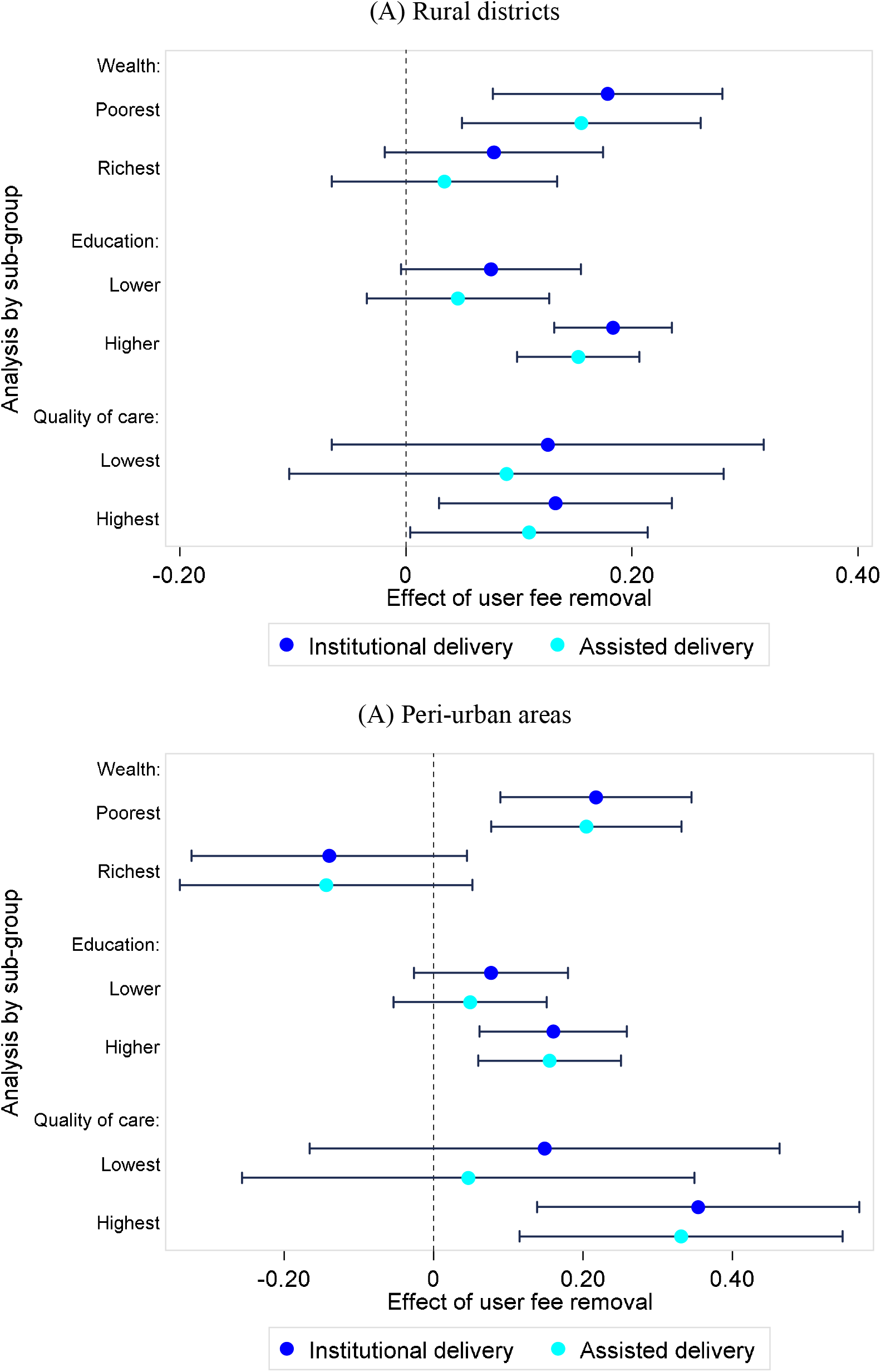
Heterogeneous effects of user fee removal.

## 4. Discussion

We analysed the effect of abolishing user fees on birth outcomes in Zambia up to six years after the policy change. Our analysis yielded four key results.

First, we found that within five to six years after the policy implementation, there was a large increase in institutional deliveries, by 25 to 35% compared to pre-reform levels. These results echo those of similar reforms where free care led to sizeable increase in institutional deliveries (Leone, Cetorelli et al. 2016, Fitzpatrick 2018). They also confirm the key role of financial barriers in accessing needed care.

A second key result was that the positive effects of the reform took time to appear. Consistent with other studies that explored the immediate effects of the policy in Zambia (Chama-Chiliba and Koch 2016, Lepine, Lagarde et al. 2018), there was no evidence of impact on maternal care-seeking up to two years after the policy change. This absence of short-term effect can be linked to implementation issues that have been documented: confusion about the definition of the policy, and lack of funding to replace lost revenue leading facilities to continue to charge fees (Carasso, Palmer et al. 2010). Our contribution is to show that positive and large effects eventually emerged, suggesting that when those implementation hurdles were overcome, behaviours changed. It is also possible that adoption of new behaviours took time to spread (i.e. delivering in a facility rather than at home). More research would be needed to tease out the relative importance of speed and quality of implementation, against behavioural change.

Our third key result was that, despite the large improvement in institutional deliveries, some groups remain left out. Our finding that women with low education did not benefit from the positive effects of the policy reform are consistent with results from other settings (McKinnon, Harper et al. 2015). This suggests that women’s lack of agency or limited information about the benefits of seeking care may still hinder care seeking and behavioural change. This calls for additional supporting interventions targeting specific group that may also be more at risk of worse health outcomes, for themselves or their new-borns.

Our last result related to the role of quality of care. On the one hand, we found that a reduction in price was not sufficient to increase the demand for institutional deliveries in areas that were plagued by the worst levels of quality of care. On the other hand, we found that despite a large increase in institutional delivery, there was no increase in the proportion of caesarean sections and no reduction in neonatal deaths. Both results point to key deficiencies in access to high quality care that is necessary to turn higher use of care into better health outcomes. While it is possible that the reform itself led to deterioration of the quality of care, for example through shortages of essential supplies or drugs (Picazo and Zhao 2009), other studies have recently underlined key deficiencies in staff skills that may require more structural reforms (Das, Woskie et al. 2018, Kruk, Gage et al. 2018).

This study has several strengths. We used nationally representative survey data to establish the systemic effects of a national reform over a long period of time. We were able to evaluate two policy changes on a range of key birth outcomes. Additionally, we were able to explore the effects of the policy for groups and in different environments, allowing us to examine the distributional effects of the policy and its potential limitations.

Some limitations of the study should be noted. Firstly, because the location of sampling clusters is randomly displaced in the DHS for confidentiality reasons, by up to 2km for urban clusters and 5km for rural clusters, we may have assigned some clusters to the wrong treatment status. Yet this problem is unlikely to be widespread, and the measurement errors and bias created by this issue are unlikely to compromise our main results. Secondly, some households in urban areas may have sought care where care was free (Lepine, Lagarde et al. 2018). This issue would have led to under-estimating the true effect of the policy. Our results are also robust to excluding three districts where such health seeking patterns were particularly prevalent (See Online Appendix E). Thirdly, the fact that caesarean sections and neonatal deaths are relatively rare events means that the study was under-powered to detect small changes. However, at least for c-sections, meaningful changes would require increases that the study should have been able to detect, but did not occur.

The study makes an important contribution to the literature on the effects of user fees, and more broadly to the current debates on how to achieve universal health coverage. Results highlight that the benefits of health financing reforms can be slow to emerge, do not always materialise for the most vulnerable populations, and will not automatically translate into better health outcomes. The concomitant introduction of supporting policies may be necessary to encourage behavioural change, especially for disadvantaged groups, for example through the provision of information or financial incentives (Powell-Jackson and Hanson 2012). Further research is needed to unpack the complex impact of health financing reforms on the provision of care, and specifically on quality of care delivered. More generally, reforms to increase access to health services should urgently seek to improve quality of care, or run the risk to waste resources.

## Data Availability

All data are publicly accessible through the DHS website

