## Supplementary material for "The long-term effects of free care on birth outcomes: Evidence from a national policy reform in Zambia": Online appendix

### A. Additional tables

##### Table A1. Effects of user fee removal (logistic regressions)

|  | **Policy change in rural districts** | |  | **Policy change in peri-urban areas** | |
| --- | --- | --- | --- | --- | --- |
|  | Odds-ratio (95%CI) | p-value |  | Odds-ratio (95%CI) | p-value |
| **Delivery in an institutional facility** | |  |  |  |  |
| Policy effect | 1·68 (1·30 to 2·18) | <0·0001 |  | 1·50 (1·07 to 2·12) | 0·019 |
| Mean pre-reform in ‘treated’ group | 0·4 |  |  | 0·38 |  |
| N | 12927 |  |  | 5126 |  |
| Pseudo R^2^ | 0·15 |  |  | 0·20 |  |
| **Assisted delivery at birth** | |  |  |  |  |
| Policy effect | 1·52 (1·17 to 1·96) | <0·0001 |  | 1·37 (0·98 to 1·91) | 0·064 |
| Mean pre-reform in ‘treated’ group | 0·39 |  |  | 0·34 |  |
| N | 12910 |  |  | 5119 |  |
| Pseudo R^2^ | 0·15 |  |  | 0·20 |  |
| **Delivery by Caesarean section** | |  |  |  |  |
| Policy effect | 1·89 (1·11 to 3·22) | 0·019 |  | 1·49 (0·65 to 3·41) | 0·570 |
| Mean pre-reform in ‘treated’ group | 0·01 |  |  | 0·02 |  |
| N | 12624 |  |  | 5108 |  |
| Pseudo R^2^ | 0·07 |  |  | 0·05 |  |
| **Neonatal deaths** |  |  |  |  |  |
| Policy effect | 1·15 (0·67 to 1·97) | 0·606 |  | 0·77 (0·32 to 1·85) | 0·566 |
| Mean pre-reform in ‘treated’ group | 0·03 |  |  | 0·03 |  |
| N | 12386 |  |  | 5120 |  |
| Pseudo R^2^ | 0·03 |  |  | 0·03 |  |

Notes: Each odds-ratio comes from a logistic regression that includes year and district fixed effects. Standard errors are clustered at mother level, sampling weights are included.

##### Table A2. Effect of user fee removal on delivery in a public or mission facility

|  | Home births | Delivery in a public or mission facility |
| --- | --- | --- |
| **Panel A: Policy change in rural districts** | |  |
| ITT effect | -0.151*** | 0.139*** |
|  | (0.020) | (0.021) |
| Pre-reform control mean | 12,965 | 0.399 |
| Observations | 0.190 | 12,927 |
| R-squared | 0.596 | 0.172 |
| **Panel B: Policy change in peri-urban areas** | |  |
| ITT effect | -0.110*** | 0.096*** |
|  | (0.033) | (0.033) |
| Pre-reform control mean | 0.618 | 0.376 |
| Observations | 5,140 | 5,126 |
| R-squared | 0.236 | 0.206 |

Notes: The table looks at the effect of removing fees on public and mission facilities. Panel A shows the impact of removing fees in rural districts, which were free from April 2006. Panel B shows the impact in peri-urban areas in urban districts, which were free from June 2017. All regressions include year and district fixed effects. Standard errors are clustered at mother level, sampling weights included,*** p<0.01, ** p<0.05, * p<0.1

##### Table A3. Heterogeneous effects of user fee removal

|  | **Rural district** | |  | **Peri-urban areas** | |
| --- | --- | --- | --- | --- | --- |
|  | Institutional delivery | Assisted institutional delivery |  | Institutional delivery | Assisted institutional delivery |
| **Panel A: Households’ wealth** | | |  |  |  |
| Poorest quintile | 0.178*** | 0.155*** |  | 0.217*** | 0.204*** |
|  | (0.052) | (0.054) |  | (0.065) | (0.065) |
| Richest quintile | 0.078 | 0.034 |  | -0.140 | -0.144 |
|  | (0.049) | (0.051) |  | (0.094) | (0.0997) |
| **Panel B: Mother’s education** | | |  |  |  |
| Low education | 0.075* | 0.046 |  | 0.077 | 0.048 |
|  | (0.040) | (0.041) |  | (0.052) | (0.052) |
| Higher education | 0.183*** | 0.152*** |  | 0.160*** | 0.155*** |
|  | (0.026) | (0.0275) |  | (0.050) | (0.0486) |
| **Panel C: Quality of care** | | |  |  |  |
| Low quality | 0.125 | 0.089 |  | 0.148 | 0.046 |
|  | (0.097) | (0.098) |  | (0.159 | (0.153) |
| High quality | 0.132** | 0.109** |  | 0.354*** | 0.331*** |
|  | (0.052) | (0.053) |  | (0.109) | (0.110) |

Notes: Each coefficient is from a single OLS regression. Panel A presents the results for the richest and poorest quintiles, determined through an asset index computed in the DHS survey. Panel B shows the results for mothers who have no primary education or who have at least primary education. Panel C shows the effect for areas offering high or low quality where high quality is defined as an area where the average quality score is the top (high quality) or the bottom (low quality) quintiles of the distribution. All regressions include year fixed effects. Standard errors are clustered at household level, sampling weights included, *** p<0.01, ** p<0.05, * p<0.1

### B. Additional Figures

##### Figure B1: Timeline of policy implementation and data sources

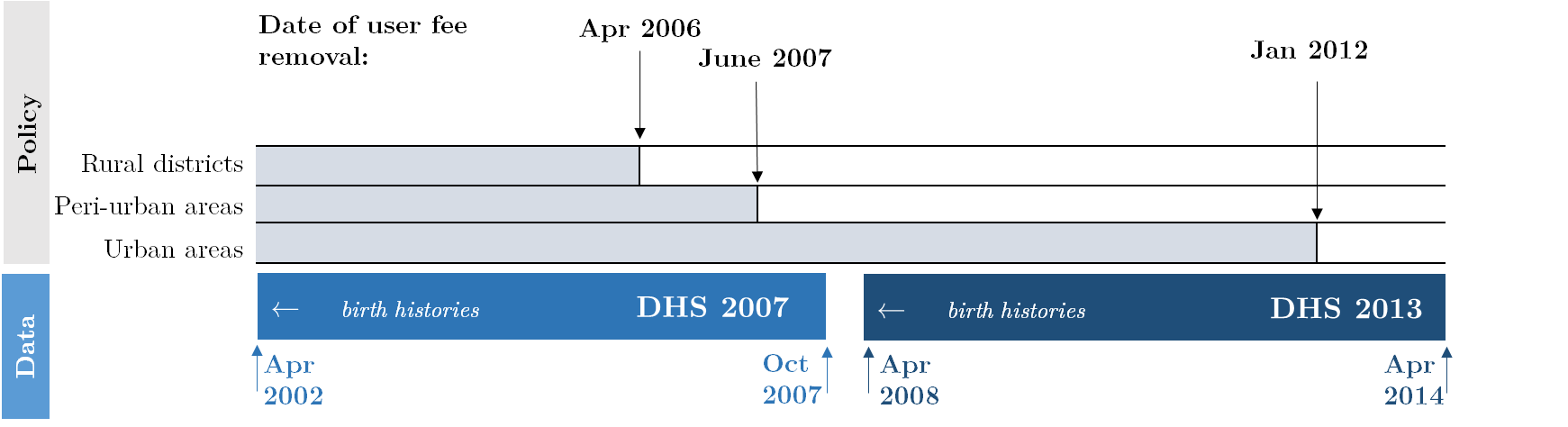

*Note*: For each DHS dataset the dates at the bottom correspond to the earliest (left) and latest (right) birth events

##### Figure B2: Trends of institutional deliveries

| (A) All institutional deliveries  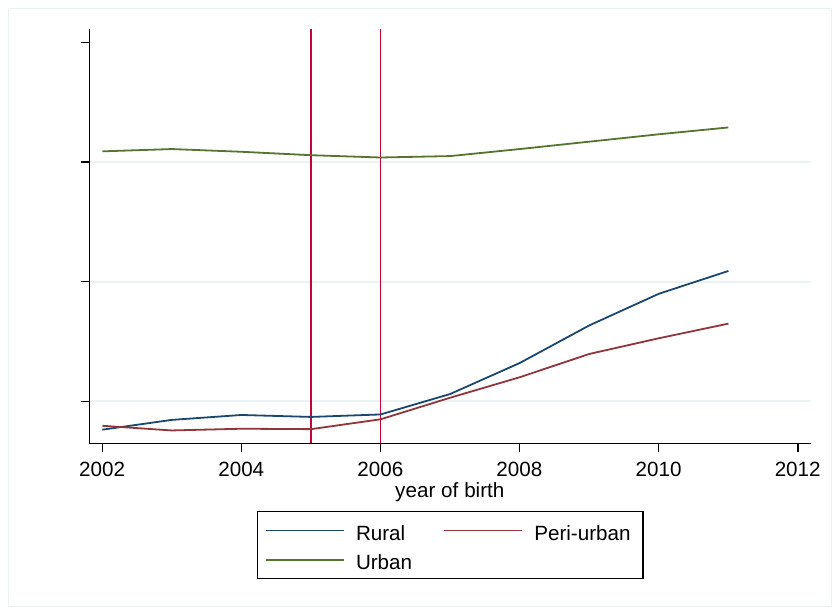 | (B) Deliveries in public and mission facilities  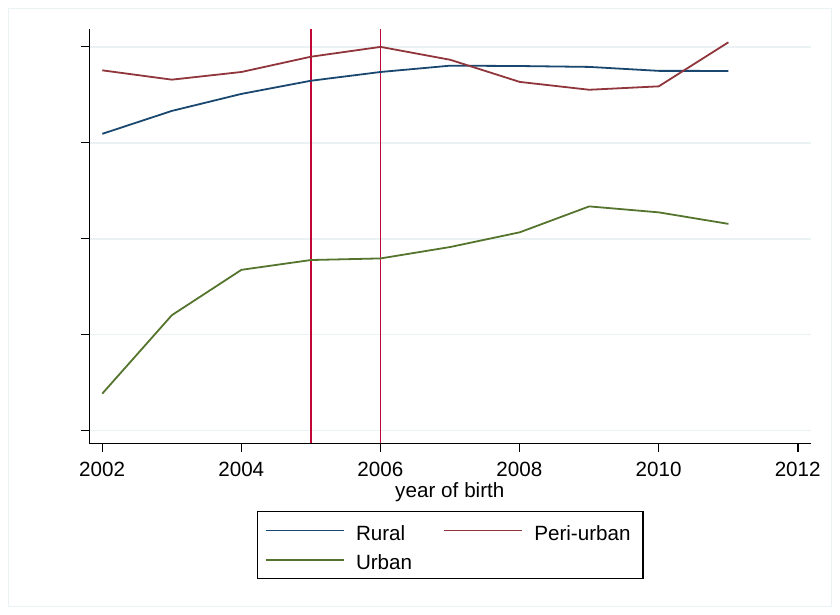 |
| --- | --- |

Note: Trends are estimated using sampling weights for the three groups. Vertical lines are drawn for the last pre-intervention year depending on the timing of the policy change (2005 for the policy change in rural areas and 2006 for the policy change in peri-urban areas).

##### Figure B3: Trend in assisted delivery at birth

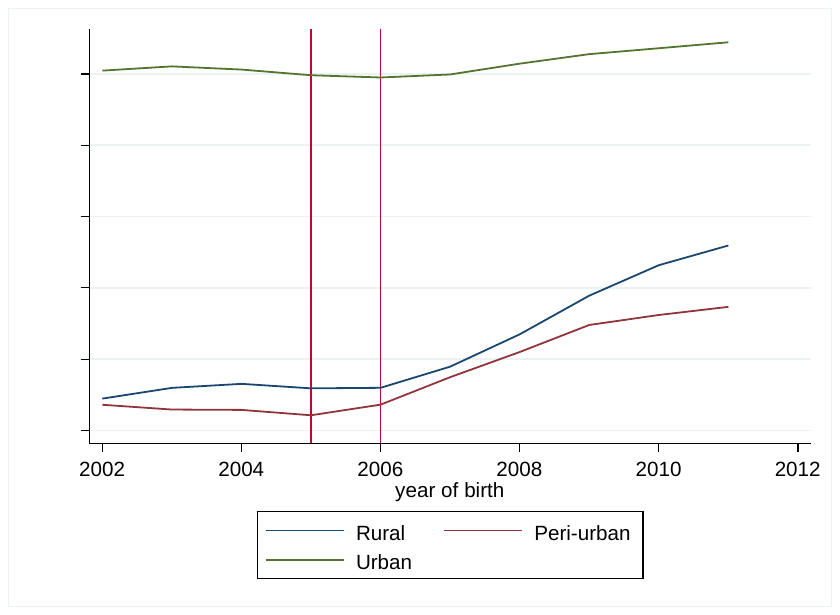

Note: Trends are estimated using sampling weights for the three groups. Vertical lines are drawn for the last pre-intervention year depending on the timing of the policy change (2005 for the policy change in rural areas and 2006 for the policy change in peri-urban areas).

##### Figure B4: Trends of the proportion of neonatal deaths

| (A) 2002-2011 | (B) 2003-2011 |
| --- | --- |
| 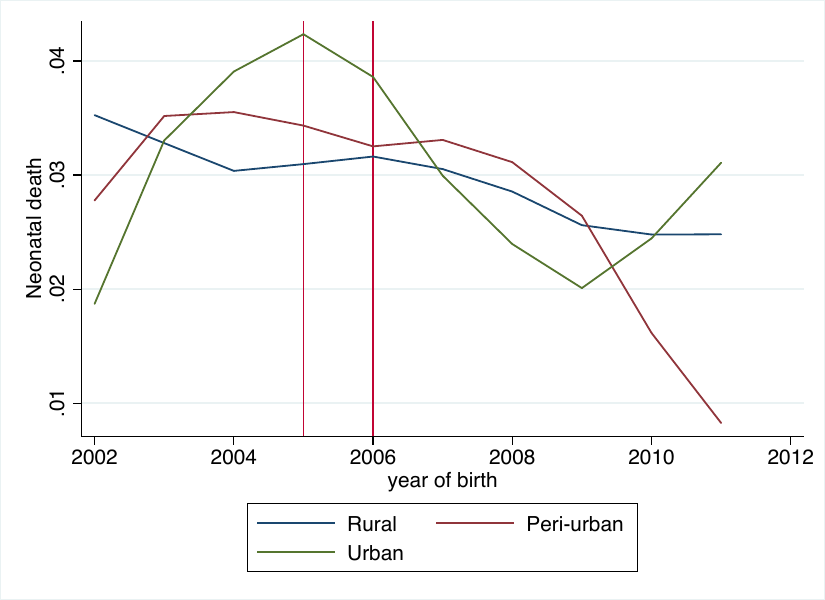 | 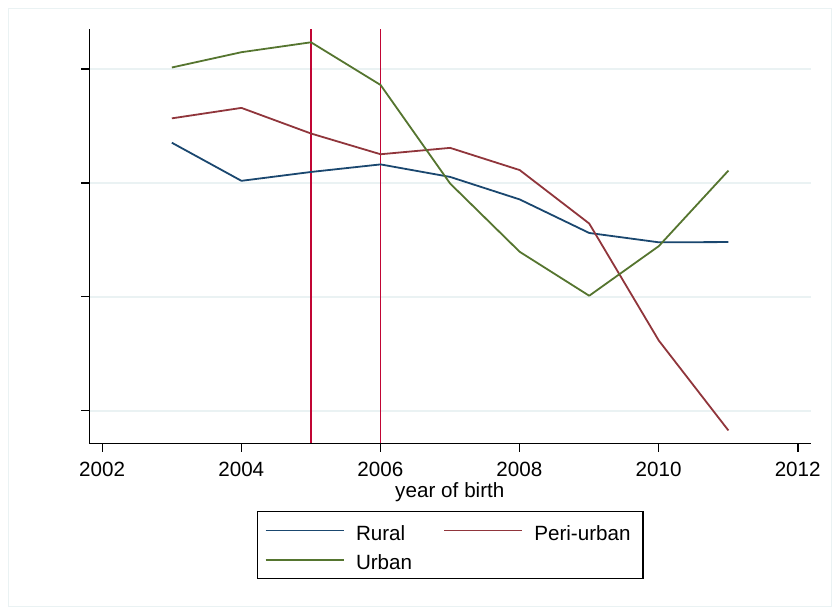 |

Note: Trends are estimated using sampling weights for the three groups. Vertical lines are drawn for the last pre-intervention year depending on the timing of the policy change (2005 for the policy change in rural areas and 2006 for the policy change in peri-urban areas).

### C. Definition of treatment and control areas

In January 2006, the Zambian president announced that user fees in health care were to be removed. The policy was to be officially implemented three months later (starting on 1^st^ of April 2006) in all primary health care facilities, health centres and district hospitals in the 54 districts designated as ‘rural’ according to the local government classification. In January 2007, user fees were removed in all public health facilities located in the peri-urban areas of the 18 districts where fees had not been abolished. The definition of peri-urban areas differed depending on whether the district was crossed by the railway or not. For districts crossed by the railway, user fees were removed for all facilities located more than 20 km away from the district administrative centre. For districts that were not crossed by the railway, and hence considered more remote, user fees were removed in all facilities located more than 15km away from the district capital. Finally, in January 2012, the free care policy was rolled out nationally, so that mission and public health facilities in urban areas of urban districts started to provide care for free (Ministry of Health, 2007). Hence, we cannot investigate the effect of the policy after 2011 due to the absence of a control group.

##### Figure C1. Distribution of DHS sampling clusters

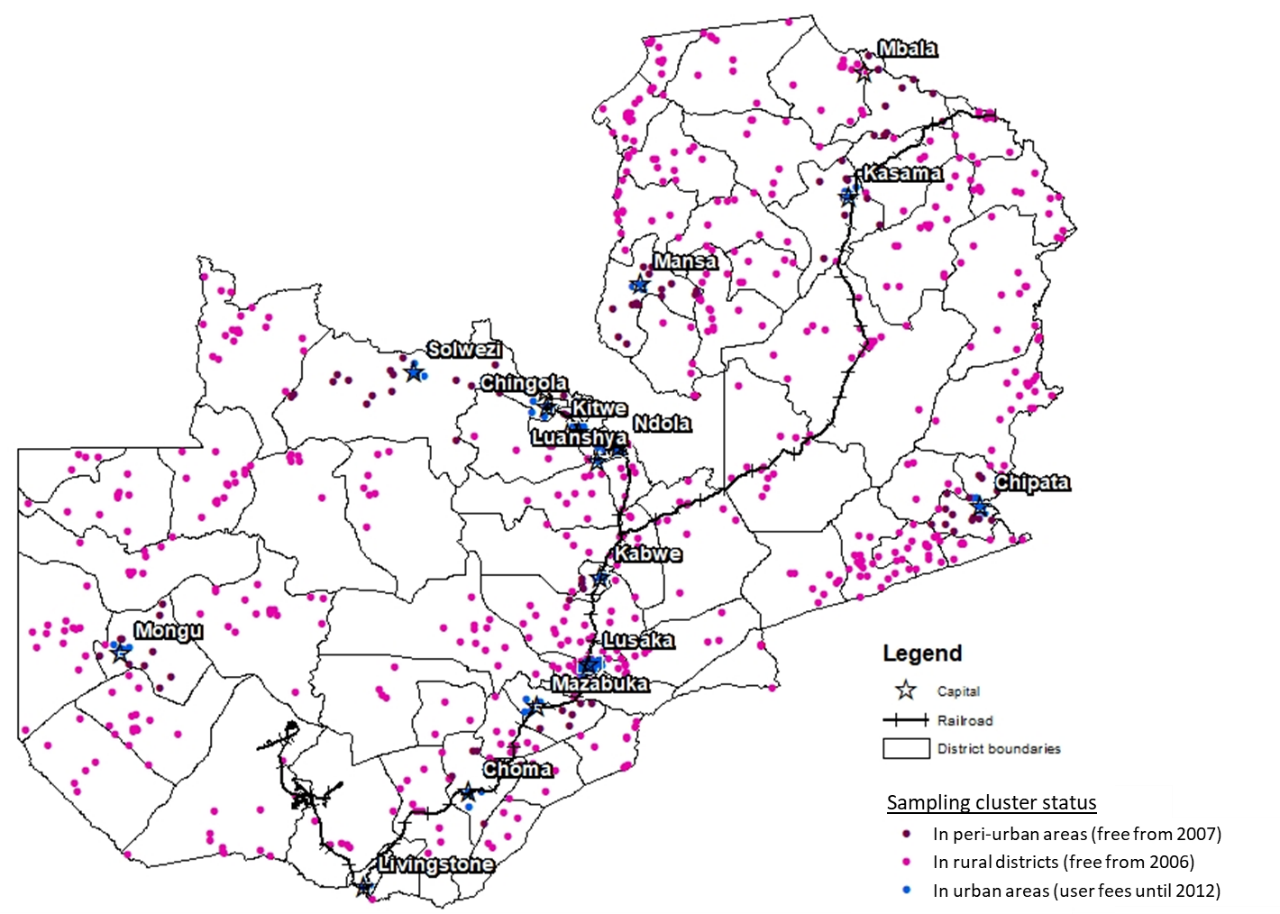

### D. Defining variables for sub-group analyses

- **Wealth**

We compute a wealth index through multiple component analysis using basic assets (television, radio, refrigerator, motorcycle/scooter, car/truck) and dwelling characteristics (access to electricity). From this indicator, we create quintiles and use the lowest and highest wealth quintiles in the sub-group analysis.

- **Quality of care**

We create a proxy indicator for the quality of care offered in an area at a certain point in time in two stages. We compute a quality index through multiple component analysis using five indicators of quality (whether the woman received vitamin at birth, was told about birth complication, had blood pressure checked, had a urine test and a blood test after delivery). Second, we compute the average quality score for all women living in the same cluster and we use the quality score for the year of birth of the child to measure the quality of care received at the time of the delivery. From this indicator, we create quintiles and use the lowest and highest quality quintiles in the sub-group analysis.

### E. Exclusion of contaminated districts

While we assume that individuals seek care in the district where they live, people from urban districts could seek care in rural districts because facilities may be closer to where they live, and obviously, after the reform, because care is free. Based on 1998 LCMS data (the only national survey for which the information is available), we find that this issue is generally limited (less than 4% of the population of urban districts seeking care in rural districts), except in three districts (Mongu, Mazabuka and Kasama) where respectively 25%, 18% and 12% of the population sought care in rural districts. To test whether this issue is partly responsible for our results, we repeated the analysis excluding those districts. The results are presented in Table E1 and show the same results as for the main analysis.

##### Table E1: Effect of user fees removal when excluding contaminated districts

|  | Institutional delivery | Institutional and assisted delivery at birth | Delivery by Caesarean section | Neonatal death |
| --- | --- | --- | --- | --- |
|  | (1) | (2) | (3) | (4) |
| **Panel A: Policy change in rural districts** | | | | |
| ITT effect | 0.141*** | 0.114*** | 0.007 | 0.009 |
|  | (0.021) | (0.022) | (0.014) | (0.010) |
| Pre-reform control mean | 0.401 | 0.391 | 0.015 | 0.029 |
| Observations | 12,345 | 12,328 | 12,364 | 12,004 |
| R-squared | 0.194 | 0.196 | 0.025 | 0.009 |
| **Panel B: Policy change in peri-urban areas** | | | | |
| ITT effect | 0.097*** | 0.079** | 0.005 | 0.008 |
|  | (0.037) | (0.036) | (0.019) | (0.013) |
| Pre-reform control mean | 0.379 | 0.341 | 0.023 | 0.03 |
| Observations | 4,191 | 4,184 | 4,193 | 4,022 |
| R-squared | 0.258 | 0.265 | 0.019 | 0.006 |

Notes: The dependent variable in column 1 takes the value 1 if the woman delivered in a health facility (public, mission and private) and 0 if she delivered at home. In column 2, the dependent variable takes the value 1 if the delivery was assisted by a professional staff (doctor, nurse or midwife). In column 3, the dependent variable takes the value 1 if the delivery was a C-section. In column 4, the dependent variable takes the value 1 if the child died within the first 28 days. Panel A shows the impact of removing fees in rural districts, which were free from April 2006. Panel B shows the impact in peri-urban areas in urban districts, which were free from June 2017. All regressions include year and district fixed effects. Standard errors are clustered at mother level, sampling weights included, *** p<0.01, ** p<0.05, * p<0.1
